# Appropriateness, barriers, and facilitators of multi-month dispensing of tuberculosis drugs in rural eastern Uganda: a qualitative study to inform a non-inferiority randomized trial

**DOI:** 10.1101/2025.04.02.25325138

**Authors:** Jonathan Izudi, Francis Bajunirwe, Adithya Cattamanchi, Nora West

## Abstract

Multi-month dispensing of tuberculosis (TB) drugs is an innovative strategy that may reduce frequent clinic visits and travel costs among people with TB (PWTB) in rural areas. To inform a planned trial, we explored the appropriateness, barriers, and facilitators to multi-month dispensing among PWTB and healthcare providers in rural eastern Uganda. We used qualitative methods situated within the Consolidated Framework for Implementation Research to explore two refill schedules for multi-month dispensing of TB treatment—a four- or five-visit refill schedule. In December 2024, we collected data through interviews with PWTB, their treatment supporters, and healthcare providers at the regional, district, and health facility levels. Data were analyzed using thematic analysis. All participants (n=39; 22 healthcare providers, 12 PWTB, and five treatment supporters) expressed willingness to adopt multi-month dispensing, with a four-visit schedule as the preferred option. Healthcare providers preferred the five-visit schedule for individuals with complex health conditions: severe illness, clinical instability, or bacteriologically confirmed pulmonary TB. Multi-month dispensing was perceived to benefit healthcare providers by reducing workload, improving patient flow, and enhancing patient management. Perceived benefits to PWTB included reduced clinic visits and travel costs, time savings, improved treatment adherence, reduced waiting times and TB-related stigma, and increased patient satisfaction with care. Facilitators included integration with existing treatment models, person-centeredness, community and family support, reliable drug supply, clear operational guidelines, healthcare provider training and readiness, enhanced monitoring and evaluation, clinic accessibility, readiness to utilize it, and leadership support. Barriers included undefined eligibility criteria, uncertain effects of multi-month dispensing, differing refill schedules for PWTB and HIV, treatment non-adherence due to forgetfulness and medication sharing, and patient disengagement due to insufficient follow-up. Multi-month dispensing is perceived to benefit PWTB and healthcare providers. Further studies to measure the impact on treatment outcomes should leverage facilitators and address barriers to adoption and effectiveness.

## Introduction

In 2022, the global treatment success rate for drug-susceptible tuberculosis (TB) was 88% [1], and 90.2% in Uganda [2]. In rural eastern Uganda, differences in treatment success rates between and within districts and health facilities are common, with most health facilities having less than 90% treatment success rate, which is suboptimal [3]. Several reasons contribute to the suboptimal treatment success rate, including frequent health facility visits and longer travel distances to TB clinics [4, 5]. Travel distances exceeding five kilometers have been associated with poorer treatment outcomes, primarily due to the financial and logistical burden of travel, which increases the risk of missed clinic visits and treatment nonadherence [3, 4].

Among people living with human immunodeficiency virus HIV (PLHIV), multi-month dispensing of anti-retroviral therapy (ART) has been shown to improve viral load suppression, mitigate stigma, reduce the risk of inadvertent HIV status disclosure, and enhance treatment adherence and retention [6–8]. These benefits may be transferable to people with tuberculosis (TB) through multi-month dispensing of TB drugs (MMD-TBD); however, empirical evidence remains limited. We hypothesized that MMD-TBD would not lead to a worse treatment success rate than routine care (bi-weekly dispensing of TB drugs for two months and then monthly for four months). To test our hypothesis, we designed the <u>M</u>ulti<u>-Mo</u>nth <u>R</u>efill of <u>A</u>nti-TB <u>D</u>rugs (MORAD) study to evaluate the preliminary effectiveness of MMD-TBD (intervention) compared to routine care in improving treatment success rate among people with drug-susceptible TB aged ≥15 years receiving the standard 6-month anti-TB regimen in rural eastern Uganda.

The MORAD study proposes two options for the MMD-TBD intervention: a four-visit or five-visit refill schedule. The four-visit option would allow monthly TB drug refills for two months, followed by a two-month refill for the next four months. The five-visit option would include bi-weekly refills for the first month, followed by a one-month refill, then a two-month refill for the remaining four months. These refill schedules would be compared with routine care, which involves eight visits: bi-weekly refills for the first two months, followed by monthly refills thereafter. However, there are uncertainties regarding the optimal refill schedule and key barriers and facilitators to MMD-TBD. To inform the randomized trial implementation, we explored the appropriateness, barriers, and facilitators to MMD-TBD among people with TB in rural eastern Uganda.

The MORAD protocol was previously published and registered with the Pan African Clinical Trials Registry (PACTR202403586718783) [9]. In this study, we utilize the abbreviation ‘MMD-TBD’ (Multi-Month Dispensing of TB drugs) to refer to providing TB drugs for multiple months. The abbreviation ‘MMD-TBD’ replaces ‘MULTI-DAT’, which was used in our previous protocol and conveys the same concept. ‘MMD-TBD’ was used instead of ‘MULTI-DAT’ to maintain clarity and consistency to prior studies that employed ‘MMD’ to mean multi-month dispensing.

## Materials and materials

### Ethical issues

The MORAD study received ethical approval from the Infectious Diseases Institute Research Ethics Committee (reference number: IDI-REC-2023-33) and the Uganda National Council for Science and Technology (reference number: HS3863ES). Administrative clearances were granted by the National TB and Leprosy Control Program at the Uganda Ministry of Health and all District Health Offices. Participants gave written informed consent after being fully informed about the study’s objectives, benefits, risks, privacy measures, confidentiality, and protections, including their right to withdraw at any time.

### Study setting and population

This qualitative study was conducted across 10 TB clinics in four districts in rural eastern Uganda. In this region, the majority of people with TB travel five or more kilometers to access TB care [3, 4]. The 10 health facilities treat 50 or more people with TB annually. Each health facility has a TB clinic managed by a TB focal person who is an experienced medical, clinical, or nursing officer. The clinics follow the National TB Control Program guidelines for standardized TB care. Participants included people with TB, their treatment supporters, and healthcare providers (HCPs) at the health facility, district, regional, and national levels. All the participants had ≥3 years of TB work experience. Eligible people with TB were those who had received TB treatment for ≥4 months, along with their respective treatment supporters. These individuals were considered to have a comprehensive understanding of TB treatment requirements based on their prior experience with TB care and were expected to provide valuable insights into the proposed MMD-TBD intervention. TB focal persons included people who had provided TB care for ≥1 year.

The HCPs were purposively selected as key informants, while people with TB were consecutively sampled. Treatment supporters were recruited using a snowball sampling method. Here, people with TB who participated in the study invited their treatment supporters to contribute insights upon request.

### Data collection and study variables

In December 2024, data were collected through key informant interviews (KIIs) with HCPs at their workplace and in-depth interviews (IDIs) with people with TB and their treatment supporters at TB clinics. S1 File shows the interview guides. The Consolidated Framework for Implementation Research (CFIR) [10, 11] guided the data collection. Details of the CFIR domains are described previously [9]. In brief, we collected data on the intervention characteristics, outer setting, inner setting, characteristics of individuals, and implementation process. Additionally, we explored the acceptability and appropriateness of the implementation strategy, including the rationale. KIIs were conducted in the English language, while IDIs were held in the local language *(“Ateso”).* All research assistants had ≥5 years of qualitative research experience and held, at a minimum, a diploma in health sciences. Field notes were taken during the data collection to capture additional details beyond the verbal responses of participants. Each interview lasted 15 to 35 minutes, on average.

### Quality control measures

We pre-tested and validated the data collection tools outside the study area to ensure reliability. Research assistants received comprehensive training on ethical research practices, the study protocol, participant recruitment, and data collection procedures. Audio recordings were transcribed within five days of collection to maintain accuracy, and a Research Team Leader oversaw the research team, reviewed transcripts, and ensured alignment with study objectives.

### Sample size and data analysis

Sample size depended on the saturation principle, a point at which no new information emerged even when additional data were collected [12]. However, the *a priori* sample size estimated to reach the saturation point was 66 participants—three participants at the Ministry of Health (MoH) level, one participant at the regional level, 12 participants at the district level, and 50 participants (10 TB focal persons, 20 persons with TB, and 20 treatment supporters) at the health facility level.

We conducted a thematic analysis to identify barriers and facilitators to implementing MMD-TBD grounded within the CFIR domains. Audio recordings were transcribed verbatim, and the emergent transcripts were verified by re-reading and replaying the audio recordings while correcting any discrepancies. Two analysts (JI and NW) familiarized themselves with the data by reading about 10 transcripts multiple times. An initial codebook was developed based on the domains, and consensus was established through discussion. The remaining transcripts were coded using inductive and deductive methods, guided by the initial codebook. JI is a public health specialist with 10 years of experience in mixed-methods research. NW is a socio-behavioral scientist with nearly 12 years of qualitative research expertise. Both JI and NW hold doctorates in public health and conducted the data analysis independently. Sub-themes were iteratively developed based on the CFIR domains, with non-representative sub-themes discarded. Two senior analysts (FB and AC) conducted the final review of the sub-themes within the CFIR domains to ensure they accurately reflected the data and study objectives. Results were reported using themes supported by illustrative participant quotes within the CFIR domains. Ethical rigor was maintained through member-checking, reflexivity to reduce subjective bias, an audit trail, and data triangulation. To ensure transparency and methodological rigor, we adhered to the Consolidated Reporting of Qualitative Studies (COREQ) guideline (S2 File) [13].

### Inclusivity in global research

Information regarding ethical, cultural, and scientific considerations specific to inclusivity in global research is included in the Supporting Information (S3 File).

## Results

### Characteristics of participants

The study involved 39 participants across four districts, as shown in Table 1. Most (59.0%) of the participants were from a health facility level, and 64.1% were male. Participants’ ages ranged from 24 to 62 years, with most (82.1%) falling within the 35–59 age category. Of the participants, 22 (56.4%) were HCPs, and 17 (77.3%) had at least 10 years of work experience.

**Table 1:** Summary of participant characteristics.

| Variables | Level | Overall (n=39) |
| --- | --- | --- |
| District | Kumi | 14 (35.9) |
|  | Ngora | 7 (17.9) |
|  | Serere | 7 (17.9) |
|  | Soroti | 11 (28.2) |
| Administrative unit | District | 15 (38.5) |
|  | Health facility | 23 (59.0) |
|  | Regional | 1 (2.6) |
| Sex | Female | 14 (35.9) |
|  | Male | 25 (64.1) |
| Age categories (years) | 24 to 34 | 6 (15.4) |
|  | 35 to 59 | 32 (82.1) |
|  | 60 and over | 1 (2.6) |
|  | mean (standard deviation, SD) | 42.0 (8.6) |
| Type of participants | HCPs | 22 (56.4) |
|  | People with TB | 12 (30.8) |
|  | Treatment supporter | 5 (12.8) |
| Highest level of education | None | 3 (7.7) |
|  | Primary | 6 (15.4) |
|  | Secondary | 6 (15.4) |
|  | Diploma | 12 (30.8) |
|  | Bachelor's degree | 9 (23.1) |
|  | Master's degree | 3 (7.7) |
| Work experience (years, n=22) | Less than 10 | 5 (22.7) |
|  | 10 and over | 17 (77.3) |
|  | mean (standard deviation, SD) | 16.6 (8.8) |
| Data collection mode | In-depth Interviews | 17 (43.6) |
|  | Key informant interviews | 22 (56.4) |

### Timing and frequency of MMD-TBD

MMD-TBD was deemed acceptable by all participants. HCPs unanimously supported a two-month TB drug refill schedule during the continuation phase, requiring only two visits. They also agreed to modify TB refills during the intensive phase to allow two or three visits. They recommended the 4-visit schedule for MMD-TBD for stable individuals—those with non-severe forms of TB and people with clinically diagnosed pulmonary TB.

> “When that patient has two visits in the first month [bi-weekly TB drug refills], and if that person is doing well, then that person can be given monthly refills to finish the initial phase. And then in the continuation phase, can now be given, every 2 months.
>
> Because that person now would have had enough counseling, would have had enough experience.” (HM #1, D1, M).

The 5-visit schedule for MMD-TBD was recommended for people with TB considered physically weak or unstable, as well as those with bacteriologically confirmed pulmonary TB. They stated that the first two weeks of the intensive phase of TB treatment would allow HCPs to assess the stability of people with TB, including monitoring for side effects and determining their readiness for MMD-TBD.

> “I think from the initial start, we give them drugs for one month. That is only if the patient, by assessment, is stable. But if the patient is not stable, that would require the patient to come back in the first two weeks again. So, I would recommend the one-month refill from the start, then ask the patient to come back after one month, then after, we give one month again to make two visits. For the continuation phase, we can give the two monthly refills.” (HCP #4, D2, M)

Furthermore, participants supported the five-visit schedule, citing the need for close monitoring of TB drug-related side effects and treatment adherence. MMD-TBD was deemed appropriate once a person with TB showed no side effects and demonstrated good adherence. An illustrative quote supporting this argument was:

> “I think the proposal is good to have five [five visits] refills in the course of [TB] treatment. Like the intensive phase, the first month of picking drugs every two weeks should remain because of the side effects of the drug on the patient and treatment adherence. So, TB treatment should not be interfered with. Then the last four months, we can give the medications twice.” (HCP #7, D3, F).

Another reason for the five-visit schedule was the need for close clinical monitoring of people with TB until their condition improved. Participants emphasized that initiating MMD-TBD should only occur once clinical improvement was achieved and consistent treatment adherence was established:

> “I agree with the schedule stating that in the intensive phase, someone [people with TB] should come every two weeks for the first month. The reason here is that for a patient who is sick with TB before we allow them to spend a month, we need to first monitor them in the first two weeks. For example, we need to know how are they responding to medicines. Whether they are getting any complications and whether they taking medicines”. (HCP #1, D4, M).

### Perceived benefits of MMD-TBD to people with TB

#### Reduced frequency of clinic visits and saves time

Among people with TB, all HCPs and people with TB reported that MMD-TBD would reduce the frequency of clinic visits and therefore lower travel costs, including saving time:

> “Every time a patient visits a health center, it is money (cost), and that money may not be there. You may find that the patients keep dodging (missing) clinic appointments because they cannot come and yet sometimes you want to refer them to the nearest facility which they may not also be willing.” (HM #1, D1, M).

#### Improves medication adherence

HCPs suggested that MMD-TBD could improve treatment adherence among people with TB, thereby contributing to optimal treatment outcomes. They noted that many individuals with TB miss clinic visits during treatment, leading to interruptions in adherence. Consequently, MMD-TBD was regarded as an innovative strategy to mitigate treatment interruptions, enhance adherence, and ultimately improve treatment outcomes.

> “It [MMD of TB drugs] will improve adherence and has been long overdue. So, this [routine care] has been causing defaulting in treatment, even adherence because sometimes some of these patients stay alone or with no caregivers. Coming to the health facility frequently will lead to somebody even abandoning their treatment.” (HCP #8, D3, F).

#### Reduces waiting times

People with TB and their treatment supporters reported experiencing long wait times at the TB clinic for drug refills. MMD-TBD was therefore considered a valuable strategy for decongesting the TB clinic and reducing patient waiting times:

> “I come here [health facility] and find the line is long [long queue] and I have to wait the whole day [longer waiting time]. And, then I have to do that again in two weeks.
>
> So, it would be better if we did [travel to the health facility] that once in a month.” (TS #1, D2, M).

#### Convenience

People with TB indicated that MMD-TBD could enhance convenience by reducing time away from work, allowing them to engage in small-scale businesses to earn a living:

> “When I am given drugs for many months [MMD of TB drugs], my business will not collapse because I will have time for it since the frequency of coming to the facility [health facility] will reduce if I am given 2 months.” (TBP #3, D1, F).

#### Minimizes TB-related stigma

People with TB often face stigma, as the disease is widely perceived as infectious and closely linked to HIV. As a result, many discontinue treatment. MMD-TBD was seen as a strategy to reduce stigma, promote treatment completion, and achieve better health outcomes.

> “This [MMD of TB drugs] will help me because some of my friends have started to tease me. They have made me fear to continue coming to collect the drugs but when it is reduced, it will save me the time and that fear”. (TBP, D2, F).

#### Increases patient satisfaction

Many HCPs indicated that the introduction of MMD-TBD would likely enhance patient satisfaction with TB care, ultimately improving overall treatment outcomes:

> “The truth is that patients will be very happy to know that this treatment [TB treatment] no longer requires frequent visits if we start giving the TB drugs every two months. They will be happy to take the treatment”. (HCP #15, D2, M)

#### Perceived benefits of MMD-TBD to HCPs Reduced workload

HCPs reported that MMD-TBD reduces workload by ensuring people with TB come to the TB clinic on appointment. They noted that this approach would help distribute patient visits more evenly throughout the week, resulting in a reduced workload:

> “I think if we come here a few times, even the health workers will not have a lot of work [reduced workload] because instead of every two weeks, patients will be coming once a month. So, once we do that, I’m sure even the health workers will not get bored of seeing us the patients.” (TS #2, D3, M).

#### Better patient management

HCPs reported that a heavy workload prevented them from providing equitable care to all patients. They further stated that the time saved through MMD-TBD could be used to provide better care to patients with greater needs:

> “We are understaffed, and having these patients visit a facility fewer times would give health workers much of the time to concentrate on the other areas of patient care.” (LFP #1, D3, M).

### Facilitators and barriers to MMD-TBD based on the CFIR domains

The emergent facilitators and barriers to MMD-TBD, categorized according to the CFIR domains, are summarized in Table 2. Our analysis identified multiple facilitators across all five CFIR domains; however, no barriers were identified within the inner and outer settings domains.

**Table 2:** Summary of facilitators and barriers to MMD-TBD based on the CFIR domains.

| CFIR domains | Facilitators | Barriers |
| --- | --- | --- |
| Intervention Characteristics | Integration with existing treatment models<br>Person-centeredness of MMD<br>Practical supporting evidence | Undefined eligibility criteria for MMD<br>Uncertain effects of MMD<br>Differing refill schedules for people with TB/HIV co-infection |
| Outer setting | Presence of community health workers for decentralized support<br>Family-level treatment support and engagement |  |
| Inner setting | • Steady availability of drugs<br>• Operational guidelines (MMD protocols and procedures)<br>• Medication instructions<br>• Enhanced monitoring and evaluation (e.g., revised data collection tools, regular reviews)<br>• Clinic accessibility for managing medication side effects and complaints |  |
| Characteristics of individuals | • HCP training and mentorships<br>• HCP readiness to implement MMD<br>• Patient motivation and readiness to utilize MMD | Non-adherence due to forgetfulness, medication sharing, and health neglect |
| Process of implementation | • Patient engagement and support (e.g., active follow-up, patient reminders, health education, and counseling)<br>• Leadership support for MMD implementation | Patient disengagement driven by insufficient follow-ups |

### Facilitators of MMD-TBD based on the CFIR domains

#### A. Intervention Characteristics

##### A1. Integration with existing treatment models

HCPs reported that MMD-TBD aligns well with existing treatment models, such as the MMD of ART and anti-hypertensives. Specifically, they noted that the MMD approach is already used for delivering ART to PLHIV and other patients, making the introduction of MMD-TBD consistent with existing MMD guidelines:

> “If you look at the schedule which is here, at least you see a patient four to five times in six months, which I think is good. And, also, I think it aligns well with the multi-month schedule for HIV, which we are doing now.” (HCP #14, D1, M).

##### A2. Person-centeredness of MMD

Participants, particularly HCPs, stated that MMD-TBD embodies a person-centered care approach by empowering individuals with TB to manage their treatment. They emphasized that its person-centered nature would facilitate/ or enable the adoption and utilization:

> “To the clients [people with TB], this approach [MMD of TB drugs] is patient-centered care so the issue of transport costs is reduced. And, the other bit also, is that we give clients a responsibility to take care of themselves away from us [the HCPs] a little.” (HCP #12, D2, M).

#### A3. Practical supporting evidence

It emerged that practical evidence supporting MMD was available, with some HCPs indicating its informal use in current practice. HCPs indicated that empirical evidence to support the use of MMD-TBD exists. They stated that based on the informal use of MMD-TBD, the approach was effective during the COVID-19 pandemic and the post-pandemic periods.

> Even I have tried [MMD of TB drugs] personally although it was not in the treatment guideline or it was not stated in any way.
>
> But we have been trying and you find the books would get rejected. You would find the dispenser trying to reduce the period. (HCP #13, D2, F).

#### B. Outer setting

##### B1. Presence of CHWs for decentralized support

The availability of community health workers (CHWs), including Village Health Team (VHT) members, was recognized as a crucial facilitator of decentralized, family-level treatment support. Participants emphasized that CHWs could also enhance adherence to directly observed therapy (DOT).

> “Another option for follow-up can be the use of the VHTs [Village Health Teams]. So, the nearby VHT can follow up on this patient and then, complies the reports and send them to the health center every month. So monthly VHT reports should include follow-up information about patients on TB treatment.” (HM, D4, M).

##### B2. Family-level treatment support and engagement

A strong family involvement in treatment adherence was highlighted as an essential facilitator for the successful uptake and implementation of MMD. Participants underscored the importance of engaging family members in supporting adherence to optimize treatment outcomes.

> “For me as a mother] who is giving the drugs to my child, I will ensure that the child takes the drugs on time. If I am not at home, I will call someone at home to ensure that the child is given the drugs on time because it will not be good for his health if he does not swallow [the TB drugs].” (TS #3, D4, F)

#### C. Inner setting

##### Steady availability of drugs

The consistent availability of TB drugs was identified by nearly all HCPs and some people with TB as the key facilitator of MMD-TBD, ensuring uninterrupted refills and supporting treatment adherence.

> “The most important thing is having the drugs available. So, for it [MMD of TB drugs] to be successful, then facilities should be stocked with enough drugs because the challenge sometimes that we face is drug stock out. So, you cannot plan to give a refill of 2 months when you have little stock. So, we must have enough stock available for us to be able to, successfully implement it.” (HCP #6, D4, M).

##### Operational guidelines (MMD protocols and procedures)

The development of operational guidelines, including standardized protocols and procedures for MMD implementation, was identified as a key facilitator for MMD-TBD. At the health facility level, participants emphasized the need for a standard operating procedure (SOP), while at the national level, they stressed that formal guidelines would be essential to support the uptake of MMD-TBD:

> “The way I know, it will be like a policy. The Minister of Health [MoH] has to produce a standard operating procedure (SOP) whereby health workers are supposed to follow. But if SOPs are not there, we cannot achieve what we want. Because in everything in health, we have SOPs and if SOPs are not there, we shall not achieve.” (TFP #1, D2, M).

##### Medication instructions

Among people with TB, clear medication instructions specifying the frequency of drug pick-ups were considered an essential facilitator for MMD-TBD by some HCPs and treatment supporters, but not by people with TB.

> “Maybe they [HCPs] can give us some write-up [medication instructions] so that we can also be reading on what you’re supposed to be doing. Because sometimes you come and you get a lot of information. And now as you are not medics [HCPs], sometimes you don’t get everything or you just get a few things.” (TS #4, D2, M).

##### Enhanced monitoring and evaluation (e.g., revised data collection tools, regular reviews)

Another key facilitator was enhancing monitoring and evaluation through updated data collection tools, such as a revised TB register that differentiates people with TB on MMD from those receiving routine care.

> “I think for those selected health facilities, the tools [data collection tools] equally need to be provided. But if we leave the tools in the way they are, we might not get the real results. So, let those columns in the TB Unit register for client visits be changed to fit MMD.” (TBP #10, D2, F)

Additionally, HCPs emphasized that regular data reviews to assess treatment outcomes are a necessary facilitator for generating evidence on MMD effectiveness. They noted that if MMD-TBD proves effective, such evidence would encourage more people with TB to adopt it.

> “There should be monthly monitoring of data to see who has not come and who has not come. Then to those who have not come, something should be done.
>
> At the monthly data review in the health center, these data should be shared or presented to all staff to know that there are patients on treatment and that new changes [MMD of TB drugs] have happened.” (HCP #22, D2, M)

##### Clinic accessibility for managing medication side effects and complaints

HCPs and people with TB acknowledged potential medication-related side effects and other health concerns among individuals initiated on MMD-TBD.

They emphasized the need for continuous access to TB clinics to ensure timely management and support, which they viewed as a key facilitator of MMD-TBD.

> “I think the health facility should constantly be open to us [accessible health facility] so that if we [people with TB] take these drugs and I develop a problem [side effects], I should be able to come back to the health facility at any time for the health workers to see me.” (TBP #17, D2, F).

#### D. Individual characteristics

##### HCP training and mentorships

The training of HCPs on MMD-TBD was identified as a crucial facilitator. Participants highlighted that the training would ensure effective implementation of MMD, including clear communication of its relevance and benefits.

> “Before this program takes place [wide implementation of MMD of TB drugs], it has been a long time since when they last trained health workers on TB. We need to organize either at the district level or at the health facility level, serious mentorships or training of the focal persons, plus even the clinical team on TB management.” (HCP #19, D2, M)

##### HCP readiness to implement MMD

The readiness of HCPs to implement MMD emerged as a key facilitator. Participants specifically highlighted the presence of TB focal persons (HCPs overseeing TB service delivery) and their high levels of readiness as crucial to the successful implementation of MMD-TBD.

> “We have no problem with it [MMD of TB drugs] because it is very good on the patient side and very good on the health worker side. At the health facility, the TB focal person is already there who is supposed to track these patients. Generate a list of patients on TB treatment at different intervals so that that list is tracked up to completion (HM, D4, M).

##### Patient motivation and readiness to utilize MMD

The readiness of people with TB to adopt MMD-TBD, demonstrated by their high motivation, was identified as a key facilitator. For example, HCPs noted that people with TB often expressed a preference for MMD-TBD but they could not help as it was not formalized.

> “Even as patients [people with TB], we have been thinking about it [MMD of TB drugs] but we have nowhere to take the idea. It has been a tough thing for us to frequently travel to the health facility to collect drugs [TB drugs]. The home is very far away and you know I have to ride a bicycle.” (TBP #14, D2, F)

Similarly, treatment supporters were receptive to MMD-TBD, noting that its implementation was long overdue.

> “I will be happy with the initiative [MMD of TB drugs] and I will even propose that they increase it to 3 months so that in a short time I will finish collecting the drugs [TB drugs].” (TS #5, D2, M)

#### E. Implementation process

##### Patient engagement and support

All HCPs indicated that the active engagement of people with TB and offering them support throughout MMD-TBD implementation would be crucial facilitators. Key modalities for achieving patient engagement and support included active follow-up by HCPs, sending reminders, and providing adequate health education and counseling. Specifically, phone calls and health education/counseling were identified as critical strategies to enhance treatment adherence.

> “We need to strengthen patient education at the facility level. Let the patient understand that at the end of 1 month, he/she is supposed to come for a refill. We need proper counseling of patients [people with TB] so that they can follow up on the treatment very well. Since we are not telling them to come often to the unit [health facility or TB clinics], they need to understand that their lives are at stake if they do not take the medicines.” (TFP #2, D3, F)

Additionally, documenting all MMD-TBD schedules to ensure easy tracking was highlighted as a key facilitator for successful implementation.

> “We [HCPs] need to do proper documentation and even put up a schedule since these patients are few. They can even display that for Patient “A”, the return date is this and for Patient “B”, the return date is that. By doing so, we make this program [MMD of TB drugs] succeed. We need to draw up a schedule of visiting clients [people with TB] at regular times based on the refills.” (HCP #18, D3, F)

##### Leadership support for MMD implementation

Participants emphasized the need to engage leaders at all levels—national, regional, district, and health facility—to ensure the effective implementation of MMD. At the district level, key stakeholders included TB focal persons (HCPs overseeing TB service delivery) and the district TB team. At the regional and national levels, engagement should include Regional TB Focal Persons (HCPs overseeing TB service delivery at the regional level) and the National TB Control Program.

> “We need to bring the leaders on board. And, when I look at the letters [support letters from NTLP support letter and the districts] that you already have here and the people being interviewed, they are people at the heart of TB management.
>
> So, the research is already giving us a direction. I feel that the same approach should be used up to the lower level so that the health facility heads [In-charges] are brought on board.” (HCP #16, D2. M).

### Barriers to MMD-TBD based on the CFIR domains

Several barriers to MMD-TBD were identified. No barriers were found within the inner and outer settings of the CFIR domains.

#### A. Intervention characteristics

##### Undefined eligibility criteria for MMD

Key barriers related to the intervention characteristics included undefined eligibility criteria for MMD-TBD. HCPs argued that MMD may not be appropriate for all people with TB.

> “All in all, giving a multi-month refill is not a bad idea, but then to which category of people are we giving a multi-month refill? I would wish that we first put a specific category of patients on monthly refills and not generalize on all. For those few or those who understand their disease, the approach would be good. As you are interacting with your patient for the first time, you can have a picture of someone who cares for themselves. For those, you can give multi-month refills if they have genuine telephone contact. Those could be the clients [people with TB] for multi-month refills.” (HCP #2, D2, F).

One criterion highlighted was to initiate MMD-TBD immediately for individuals with clinically diagnosed pulmonary TB but to delay it by one month for those with bacteriologically confirmed pulmonary TB.

> “We should start multi-month for PCDs [persons with pulmonary clinically diagnosed TB] and we keep the PBCs [persons with bacteriologically confirmed pulmonary TB] at the health facility because they are very infectious. We want to retain them to ensure that they take their treatment correctly and also get enough information to make them understand that they need to adhere to their treatment and take it for their full duration.” (HCP #15, D3, M).

##### Uncertain effect of MMD

HCPs expressed concerns about the unknown effect of MMD-TBD in improving treatment adherence and outcomes compared to routine care. Consequently, they called for a randomized trial before its implementation.

> “You need to evaluate it. Is it doable? Is it helping? You need to make a comparison between the new approach and the present one [MMD of TB drugs]. You need to follow it up to see if it is working [asses effectiveness]. Of course, whether it is affecting the TB outcome and what the treatment outcomes are.
>
> So, it is just a matter of getting the data comparing the previous [routine care] and the current [MMD of TB drugs] to see if it is beneficial.” (HCP #13, D2, F).

##### Differing refill schedules for people with TB/HIV

For people with TB and HIV, differences in refill schedules for ART and TB drugs emerged as an additional barrier; however, this was considered manageable, as both MMD-TBD and ART can be aligned.

> “What is again important is we have to harmonize TB and HIV refills so that the pill burden and the appointment date match. Because, you know, most ARVs, are packed for three months and one month. And, most of the health facilities I have visited have ARVs packed for three months. We can divide the pills, but again, dividing it and how to keep the balance is a challenge.” (TFP #1, D2, M)

#### B. Characteristics of individuals

##### Non-adherence

###### a. High pill burden

Treatment nonadherence emerged as a significant barrier within the CFIR construct of individual characteristics. Major factors contributing to nonadherence included perceived treatment burden, as people with TB may reduce medication intake due to a high pill burden.

> “Outside the health facility, the patient may mean to relax when gets too many drugs at ago. The patient would say can’t this be enough really to cure this [TB disease]? The patient now starts negotiating within himself or herself.” (HCP #13, D2, F).

###### b. Alcohol consumption

Concerns were raised by some HCPs about alcohol consumption among people with TB enrolled on MMD-TBD. Locally brewed alcohol is widely sold in the rural eastern region as an alternative source of livelihood, and many people with TB consume it. This could hinder treatment adherence, as the MMD-TBD approach reduces the need for frequent visits to health facilities.

“One of the worst things that might affect [MMD of TB drugs] will be alcohol drinking because somebody will even forget to take drugs.” (LFP #1, D3, M).

However, some HCPs and treatment supporters suggested that health education or counseling on the dangers of alcohol consumption could help reduce alcohol use. Some illustrative quotes supporting this claim include the following:

> “With such people [alcoholics], I think if the program is to work better, we just need information [health education or counseling]. We give them more information, encourage them, and let them know that we are proposing to reduce the number of times for going to the hospital to collect drugs.” (TS #6, D4, M).

###### c. Forgetfulness

A few HCPs suggested that the MMD-TBD approach could lead to treatment non-adherence, as people with TB might forget to take their pills. They indicated that, since the pills would be available for a longer period, it may not be obvious to people with TB that they need a refill.

> “There’s a tendency to forget to take the drugs once you have all of them [TB drugs]. You will forget to swallow them.” (HCP #7, D4, F).

###### d. Medication sharing

Participants noted that medication sharing among people with TB exists, although it is not very common. Particularly, they said that the tendency to share medications arises when patients receive refills that last for longer periods.

> “We have also realized that when some patients are given more drugs, they start to share them with other people or use them for other things because the duration of refill is not calling for their immediate return to the clinic.” (HCP #12, D1, F).

###### e. Health neglect

Some people with TB suggested that a few of their colleagues neglect their health by not taking their medications, even when refilled for longer periods. They proposed that ongoing counseling and encouragement could help reduce this neglect.

> “Someone may not take the drugs once given. This is because some people are careless and do not love themselves. But when we have someone at home to keep advising and encouraging the person, the person will take it.” (TBP #9, D1, F).

#### C. Implementation process

##### Patient disengagement driven by insufficient follow-ups

Within the implementation process, patient disengagement due to insufficient follow-up was identified as a barrier. Participants noted that inadequate follow-up for people on MMD-TBD could result in more frequent loss-to-follow-up, worsening treatment outcomes.

> “This [MMD] needs some careful implementation and follow-up so that it does not lead us to high loss to follow-up and [treatment] failure rates because, without the proper follow-up of these patients, they may come back when they are worse than the way we started [on treatment].” (HM, D1, M).

## Discussion

This qualitative study was designed to inform a randomized trial on refill scheduling and assess the appropriateness, barriers, and facilitators of implementing MMD-TBD in rural eastern Uganda. The findings revealed a consensus among participants on the need to adopt MMD-TBD. The four-visit schedule was the most preferred option while the five-visit schedule was recommended by HCPs for people with complex health conditions, including those who are severely ill, clinically unstable, or diagnosed with bacteriologically confirmed pulmonary TB. These data support further evaluation of MMD-TBD as a person-centered strategy to improve TB treatment outcomes. Longer TB drug refills are increasingly being considered a best practice to ensure uninterrupted treatment, even for patients requiring more frequent clinic visits due to clinical disease severity. For instance, a 2-month TB drug refill during the intensive treatment phase has been proposed to support treatment completion [14].

Our study showed that MMD-TBD is perceived to offer several benefits for HCPs (including reducing workload, saving time, and enhancing patient management) and people with TB (including saving time, reducing travel costs, increasing convenience, minimizing stigma, and improving treatment adherence, and satisfaction). To the best of our knowledge, no published study has focused on MMD-TBD. However, our findings are consistent with several studies of MMD of ART conducted among people living with HIV. These studies have shown that MMD of ART decongests health facilities, reduces HCP workload, alleviates HCP work burnout [15], and improves health service delivery and ART adherence [16]. MMD of ART for up to 6 months has also been shown to significantly reduce HIV clinic volumes and enhance access to treatment by improving care continuity and reducing patient travel burden [17]. Similarly, MMD-TBD may help address health system challenges and alleviate the burden on affected individuals and families [18].

We found several facilitators of MMD-TBD, including its alignment with existing treatment models and a person-centered care approach. These facilitators resonate with findings from studies on anti-hypertensives, which also highlight patient-centered care as beneficial for stable individuals [19]. Findings around HCP training, data review, HCP readiness to start MMD-TBD, and patient engagement and support systems such as active follow-up, reminders, health education, and counseling are consistent with a previous study on a six-month MMD of ART implementation [20]. We found support from community health workers (CHWs) and family members facilitates treatment adherence among people with TB on MMD, which aligns with previous studies in rural eastern Uganda highlighting a strong social support system, including treatment supporters, for achieving optimal treatment success [3, 21]. HCP training and their readiness to implement MMD-TBD, along with the willingness of people with TB to engage in MMD-TBD, were identified as key facilitators. These findings are consistent with a study among PLHIV, which reported an increasing demand for MMD as a significant facilitator [22].

Leadership support emerged as a crucial factor in facilitating MMD-TBD implementation which is consistent with the pivotal role of leadership in successfully expanding access to TB drugs through MMD through PEPFAR’s partnership with ministries of health during the COVID-19 pandemic response [23]. Additionally, operational guidelines and enhanced monitoring, along with revised data collection tools and regular reviews, are key facilitators for implementing MMD-TBD. While previous studies do not directly support these findings, effective data collection and analysis are crucial for evaluation. Tools like TB unit registers enable accurate documentation and regular reviews aid progress tracking. These have improved the uptake of MMD of ART in Tanzania [22].

This study identified several barriers to MMD-TBD, including unclear eligibility criteria, uncertain effects, mismatched refill schedules for people with TB and HIV, and adherence challenges such as pill burden, alcohol use, forgetfulness, medication sharing, and health neglect. These barriers align with findings from previous studies involving PLHIV, which noted challenges related to medication storage and infrequent health monitoring [15]. While improved drug availability is essential, risks such as stockouts and logistical issues like missed appointments due to misaligned dispensing and testing schedules also pose challenges [16, 20].

HCPs identified medication sharing and storage as barriers; however, people with TB and their treatment supporters reported no such concerns, highlighting a discrepancy between perspectives [18]. Barriers such as patient disengagement primarily from inadequate follow-up, and treatment non-adherence significantly impact treatment outcomes among people with TB in low-resource settings [24] but can be mitigated by MMD-TBD through reduced clinic visit frequency, lowered transportation costs, and enhanced treatment adherence.

### Strengths and limitations

Our study has several strengths. Our study is the first to propose MMD-TBD for people with TB in rural Uganda and potentially sub-Saharan Africa. Data collection and analysis were grounded in a well-established theoretical framework, the CFIR. This may result in a theory-informed intervention with high acceptability, appropriateness, and scalability. We gathered data from diverse stakeholders involved in TB care delivery at the regional, district, and health facility levels, ensuring that the findings are both context-relevant and policy-aligned. Additionally, data were collected from people with TB, the direct beneficiaries of MMD-TBD, and their treatment supporters, further enhancing the acceptability and applicability of the findings. Saturation was reached during data collection, ensuring adequate depth for drawing valid conclusions.

Despite the study’s strengths, there are limitations. The generalizability of findings to other contexts may be constrained due to differences in social, economic, policy, and environmental factors. The richness of the data depended on the expertise and openness of the participants, which may have been influenced by social desirability bias.

To address these limitations, experienced qualitative research assistants employed rigorous probing techniques, ensuring comprehensive and reliable insights. Concerns about undefined eligibility criteria for MMD-TBD will be addressed by aligning intervention delivery with the elements of differentiated service delivery (DSD) models in Uganda. This approach will tailor MMD-TBD to the clinical characteristics of people with TB—whether stable, unstable, or complex—and to specific populations such as adults, children, and adolescents, as well as contextual factors like rural or urban settings and stability of the environment. The uncertain effect of MMD among HCPs compared to routine care will be addressed through a forthcoming noninferiority randomized trial. The trial will align with Uganda’s DSD guidelines for TB and HIV, which emphasize the need for evidence to support differentiated care. As the next step, we will conduct a trial to compare the effectiveness of MMD-TBD with routine care. The 4-visit schedule of MMD-TBD will be assigned to stable people with TB, while the 5-visit schedule will be assigned to those considered unstable, very sick, or diagnosed with bacteriologically confirmed pulmonary TB.

### Conclusion and recommendations

The study findings demonstrate that MMD-TBD is a feasible and beneficial approach for both HCPs and people with TB. The most preferred MMD option was a 4-visit schedule, with a 5-visit schedule preferred for people with complex health conditions. MMD-TBD not only reduces HCP’s workload but also offers significant advantages for people with TB, including reduced travel costs, time savings, increased convenience, reduced stigma, and improved treatment adherence. Facilitators included integration with existing treatment models, CHW support (along with family-level treatment support), steady drug availability, and the readiness of HCPs and people with TB for MMD-TBD implementation.

Barriers included undefined eligibility criteria, uncertain effects of MMD, and differences in ART and TB drug refill schedules among people with TB/HIV should be addressed to maximize the effectiveness of MMD. Further studies to measure the impact of MMD-TBD on treatment outcomes should leverage facilitators and address barriers to adoption and effectiveness.

## Data Availability

All relevant data are within the paper.

## Acknowledgments

We gratefully acknowledge the Center for Effective Global Action (CEGA) at the University of California for their mentorship support of the primary author. Our sincere thanks also go to the research assistants for their dedicated efforts in data collection. We express our appreciation to the District Health Officers in Soroti, Kumi, Serere, and Ngora for their invaluable administrative support. Finally, we extend our gratitude to all study participants for their essential contributions to the MORAD study.

## Supporting information

S1 File: Data Collection Instruments.

S2 File: Consolidated Reporting of Qualitative Studies (COREQ) guideline.

S3 File. Inclusivity in global health research.

